# An emergent, high-fatality lung disease in systemic juvenile arthritis

**DOI:** 10.1101/19002923

**Authors:** Vivian E. Saper, Guangbo Chen, Gail H. Deutsch, R Paul. Guillerman, Johannes Birgmeier, Karthik Jagadeesh, Scott Canna, Grant Schulert, Robin Deterding, Jianpeng Xu, Ann N. Leung, Layla Bouzoubaa, Khalid Abulaban, Kevin Baszis, Edward M. Behrens, James Birmingham, Alicia Casey, Michal Cidon, Randy Cron, Aliva De, Fabrizio De Benedetti, Ian Ferguson, Martha P. Fishman, Steven I. Goodman, Brent Graham, Alexei Grom, Kathleen Haines, Melissa Hazen, Lauren A. Henderson, Assunta Ho, Maria Ibarra, CJ Inman, Rita Jerath, Khulood Walid Khawaja, Daniel J Kingsbury, Marisa Klein-Gitelman, Khan Lai, Sivia Lapidus, Clara Lin, Jenny Lin, Deborah R. Liptzin, Diana Milojevic, Joy Mombourquette, Karen Onel, Seza Ozen, Maria Perez, Kathryn Phillippi, Sampath Prahalad, Suhas Radhakrishna, Adam Reinhardt, Mona Riskalla, Natalie Rosenwasser, Johannes Roth, Rayfel Schneider, Dieneke Schonenberg-Meinema, Susan Shenoi, Judith A Smith, Hafize Emine Sonmez, Matthew L. Stoll, Christopher Towe, Sara O. Vargas, Richard K Vehe, Lisa R. Young, Jacqueline Yang, Tushar Desai, Raymond Balise, Ying Lu, Lu Tian, Gil Bejerano, Mark M. Davis, Purvesh Khatri, Elizabeth D. Mellins, the Childhood Arthritis and Rheumatology Research Alliance Registry Investigators

## Abstract

**Objective:** To investigate characteristics and risk factors of a novel parenchymal lung disease, increasingly detected in systemic juvenile idiopathic arthritis (sJIA).

**Methods:** In a multi-center retrospective study, 61 cases were investigated, using physician-reported clinical information and centralized analyses of radiologic, pathologic and genetic data.

**Results:** Lung disease (LD) was associated with distinctive features, including acute erythematous clubbing and a high frequency of anaphylactic reactions to the IL-6 inhibitor, tocilizumab. Serum ferritin elevation and/or significant lymphopenia preceded LD detection. The most prevalent chest CT pattern was septal thickening, involving the periphery of multiple lobes +/- ground glass opacities. Predominant pathology (23/36) was pulmonary alveolar proteinosis and/or endogenous lipoid pneumonia (PAP/ELP), with atypical features, including regional involvement and concomitant vascular changes. Apparent severe delayed drug hypersensitivity occurred in some cases. 5-year survival was 42%. Whole-exome sequencing (20/61) did not identify a novel monogenic defect PAP-related or macrophage activation syndrome (MAS)-related mutations as likely primary cause. Trisomy 21 (T21) increased LD risk, as did young sJIA onset. Refractory sJIA was not required for LD development. Exposure to interleukin (IL)-1 and IL-6 inhibitors (46/61) was associated with multiple LD features. By several indicators, severity of sJIA was comparable in drug-exposed subjects and published sJIA cohorts. MAS at sJIA onset was increased in the drug-exposed, but it was not associated with LD features.

**Conclusions:** A rare, life-threatening LD in sJIA is defined by a constellation of unusual clinical characteristics. The pathology, a PAP/ELP variant, suggests macrophage dysfunction. Inhibitor exposure may promote LD, independent of sJIA severity, in a small subset of treated patients. Treatment/prevention strategies are needed.

## INTRODUCTION

Systemic juvenile idiopathic arthritis (sJIA) is a chronic, inflammatory disease of childhood, observed worldwide, with an incidence of 0.4-0.9/100,000 in North America and Europe.[1] A similar disease occurs in adults [adult-onset Still’s disease (AOSD); incidence (0.2-0.4/100,000)].[2] sJIA is characterized by a combination of arthritis, which can be destructive, and systemic inflammation, including daily fever spikes, evanescent macular rash, and serositis. A life-threatening complication (mortality rate: 8-17% in sJIA)[3] is overt macrophage activation syndrome (MAS). MAS is a form of secondary hemophagocytic lymphohistiocytosis (HLH) that manifests as a cytokine storm, with very high serum ferritin and, in severe cases, organ failure. Therapies that antagonize cytokines IL-1 and IL-6 were introduced into management of sJIA ∼15 years ago. This approach is rapidly effective in >65% patients, implicating these inflammatory mediators as key drivers of sJIA.[4-6]

The usual pulmonary complications of sJIA are pleuritis and pleural effusion.[1,7] Scattered case reports of other lung diseases in sJIA have appeared.[8-12] However, in the last decade, pediatric rheumatologists have increasingly detected cases with types of lung diseases seen only rarely in sJIA previously. Kimura *et al* described 25 cases that occurred before February 2011; diagnoses included pulmonary arterial hypertension (64%), interstitial lung disease (28%), and pulmonary alveolar proteinosis (20%). sJIA course was considered severe, with MAS in 80%.[13]

Here, we performed a multi-center, retrospective study of 61 cases, with centralized analyses of radiographic, pathologic and genetic data to provide a current characterization of lung disease in sJIA or sJIA-like disease, search for early indicators and investigate risk factors.

## METHODS

### Case definition and comparators

We used an operational sJIA case definition, developed by expert consensus as a modification of the ILAR (International League of Associations for Rheumatology) sJIA classification criteria.[14] Cases failing to meet the case definition, but managed clinically like sJIA, were classified as sJIA-like. We identified patients with sJIA or sJIA-like disease and parenchymal lung disease (LD) through the Childhood Arthritis and Rheumatology Research Alliance (CARRA) network and the international pediatric rheumatology listserv (administered by McMaster University, Ontario). Inclusion required verification of sJIA or sJIA-like illness, data at LD diagnosis, and parenchymal LD by chest CT and/or lung tissue, available for expert review. Comparator data were from physician-classified sJIA patients without known LD, enrolled in the CARRA registry (CR)[15] between 2015-2018, or in the PharmaChild pharmacovigilance registry.[16] See **figures S1A-B, S2** and online supplementary information for additional details.

### Data collection

Institutional Review Board permission was obtained at each institution according to local requirements. Data included demographics, medical history, clinical features at sJIA onset, at LD diagnosis, and at defined time points (visits) before and after LD diagnosis. RegiSCAR score for drug related eosinophilic systemic syndrome (DReSS) was calculated from clinical data.[17] Online supplementary information includes details and definitions (**table S1**).

### Data analysis

Centralized analyses were performed for chest CT scans from 58/61 subjects, histopathology from 36/61 subjects and whole exome sequence (WES) data from 20 subjects. R program, version 3.5.1, was used for all statistical analyses. Additional methods are in online supplementary information.

## RESULTS

### Disease characterization: features unusual for sJIA

The cohort included 45 cases of sJIA and 16 cases of sJIA-like disease; findings prior to or associated with LD did not differ systematically between these two groups (**tables S2-4**). Demographics of the LD cohort are mostly similar to sJIA subjects in the CARRA registry (CR), with the notable exceptions of significantly lower median age at sJIA onset and significantly higher prevalence of trisomy 21 (**table 1**). These findings are discussed further below (*Risk factors*).

**Table 1:** Demographic characteristics.

| **Table 1: Demographic characteristics** |  |  |  |
| --- | --- | --- | --- |
|  | **sJIA-lung disease cohort** | **sJIA (CR)** | **P value<sup>1</sup>** |
| Sex (Female %) | 66% (40/61) | 59% (278/471) | 0.41 |
| Age [median years (IQR)] | 2.8 (1.2 - 6.3) | 5.2 (2.8 - 9.8) | 1.7 x 10<sup>-5</sup> *** |
| Race |  |  | 0.53 |
| White | 62% (38/61) | 66% (308/466) |  |
| Black | 8.2% (5/61) | 11% (50/466) |  |
| Other<sup>2</sup> | 30% (18/61) | 23% (108/466) |  |
| Region (Only U.S.)<sup>3</sup> |  |  | 0.02 * |
| Northeast | 19% (10/53) | 28% (123/442) |  |
| Midwest | 25% (13/53) | 29% (129/442) |  |
| South | 17% (9/53) | 23% (101/442) |  |
| West | 40% (21/53) | 20% (89/442) |  |
| Genetics |  |  |  |
| Trisomy 21 | 9.8% (6/61) | 0.2% (1/471) | 0.04*<sup>4</sup> |
| Familial HLH | 7.1% (2/28)<sup>5</sup> | - | - |
| *: p < 0.05; **: p < 0.01; ***: p < 0.001 |  |  |  |
| <sup>1</sup>For categorical items, Fisher's exact tests (including the multi-category form) were performed. For age, a Wilcoxon rank-sum test was performed; inter-quartile range, IQR |  |  |  |
| <sup>2</sup>Other: Hispanic 5; Middle Eastern 1; Asian 2: multi-ethnic 8; other 2 |  |  |  |
| <sup>3</sup>Based on US Census Bureau population-balanced regions: www2.census.gov/geo/pdfs/maps-data/maps/reference/us_regdiv.pdf |  |  |  |
| <sup>4</sup>Odds ratio (OR)=50 |  |  |  |
| <sup>5</sup>Diagnosed by clinical testing; one with 2 UNC13D mutations; one with UNC13D/PRF1 mutations; both sJIA-like. |  |  |  |
| CR, CARRA registry |  |  |  |

Clinical features prior to LD and associated with LD are summarized on **tables S2** and **S3**, respectively. At LD diagnosis, respiratory signs and symptoms were typically absent or subtle, although hypoxia was reported in 43% and clinical pulmonary hypertension (PH) in 30%. Strikingly, 61% of patients developed acute clubbing, sometimes as the first indicator of LD. In over half of these, digital erythema occurred (**figure 1A-C**). Other atypical features were pruritic, non-evanescent rashes in 56% (**figure 1D-F)**, eosinophilia in 37%, unexplained, severe abdominal pain in 16% (likely underestimated as this was not directly queried). Anaphylaxis to tocilizumab was unusually common, occurring in 38% of those exposed (14/37), compared to 0.6% (1/159) in the CR, 0.9% (1/110) in the tocilizumab trial in sJIA.[5a]. In one study, severe tocilizumab reactions occurred in 3/11 young (<2 years old) active sJIA patients, without known lung disease [5b]; however, in the LD cohort, the median age at reactions was 4.15 years (IQR:2.58y-6.04y). Overall, the LD cohort manifested clinical features that are unusual for sJIA [18] or pulmonary disease.

**Figure 1:**
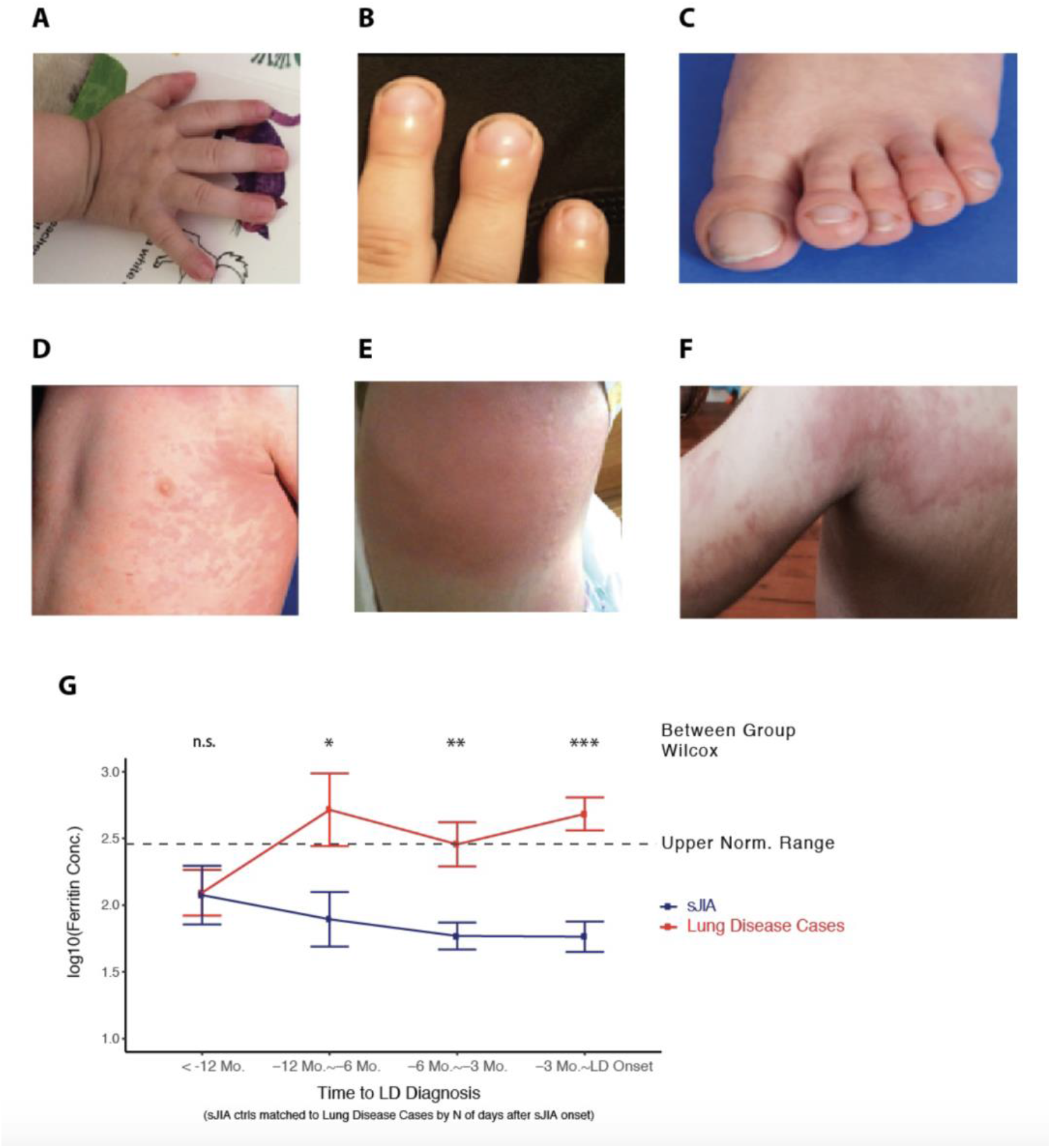
Distinctive clinical features in sJIA with lung disease (LD) and survival outcome. A. Acute erythematous digital clubbing. B, C. Bulbous deformity with erythematous clubbing of fingers (B), toes (C). D. Typical salmon-colored, macular sJIA rash (evanescent). E, F: Atypical rashes that occur before lung disease detection. E. Edematous, urticarial, non-evanescent rash (knee). F. Serpiginous, eczematous, non-evanescent rash with hyperpigmented borders. G. Mean ± standard error blood ferritin values of propensity matched (**figure S3**) sJIA controls (blue) and lung disease cases (red) across time points relative to LD diagnosis. n.s., P> 0.1; *, P < 0.05; **, P < 0.01; ***, P < 0.001, by Wilcoxon ranking sum test.

### Candidate early indicators of lung disease in sJIA: ferritin and lymphopenia

Median time to LD diagnosis after sJIA onset was 1.6 years (IQR 0.8y-3.3y), excluding 6/61 cases with LD at systemic disease onset. To identify candidate early signs of lung disease, we analyzed laboratory values commonly followed in sJIA. We matched cases to sJIA controls from the CR (1:1) for factors including laboratory test timing relative to sJIA onset, overall drug exposure, sex, sJIA onset age (**figure S3**). We then assessed serum ferritin as an indicator of inflammation. Mean serum ferritin level in cases one year prior to LD diagnosis was not distinguishable from that of propensity matched sJIA patients in the CR. However, the level rose substantially within the 12 months before LD diagnosis in the cohort (**figure 1G, figure S4**).

Another finding that preceded LD detection was significant lymphopenia [absolute lymphocyte count (ALC) <60% of age-adjusted, lower limit of normal].[19] This degree of lymphopenia, without concurrent MAS, was documented between the 6 month and 1 month visit prior to LD diagnosis in 42% of cases (excluding those with LD at sJIA onset; **table S3**). We were unable to compare this to CR controls due to lack of information. However, this degree of lymphopenia is not a known feature of active sJIA.[18] Increased ferritin and lymphopenia before LD diagnosis suggest a possibly extended incubation phase associated with smoldering inflammation and/or delayed recognition of lung disease.

### Radiologic features

As a step towards determining the nature of the LD, chest CT scans from 58/61 patients, most obtained at diagnosis, were systematically reviewed (RPG). Most exhibited one or more of 5 patterns **(figure 2A-E**). Pattern A (septal thickening involving the periphery of multiple lobes, most marked in the lower lung zones, parahilar/paramediastinal and/or anterior upper lobes with or without adjacent ground-glass opacities) was the most frequently observed (60%). Crazy-paving (B), peripheral consolidation (C), peribronchovascular consolidation (D), and predominantly ground-glass opacities (E) were seen in 21%, 22%, 16%, and 12% respectively. Among those with contrast-enhanced CTs, 11/30 (37%) displayed hyper-enhancing lymph nodes (**figure 2F**), a peculiar finding, previously reported in unusual conditions.[20,21] Findings like pattern A have been observed with connective tissue disease (CTD)-associated interstitial lung disease or interstitial pneumonia with autoimmune features.[22]. However, unlike these disorders, radiologic signs of fibrosis (honeycombing, traction bronchiectasis) were uncommon in our cohort. Overall, the observed CT findings are unexpected in sJIA; the more typical finding of pleural effusion[1,7,18] was rare at LD diagnosis (**table S4**).

**Figure 2:**
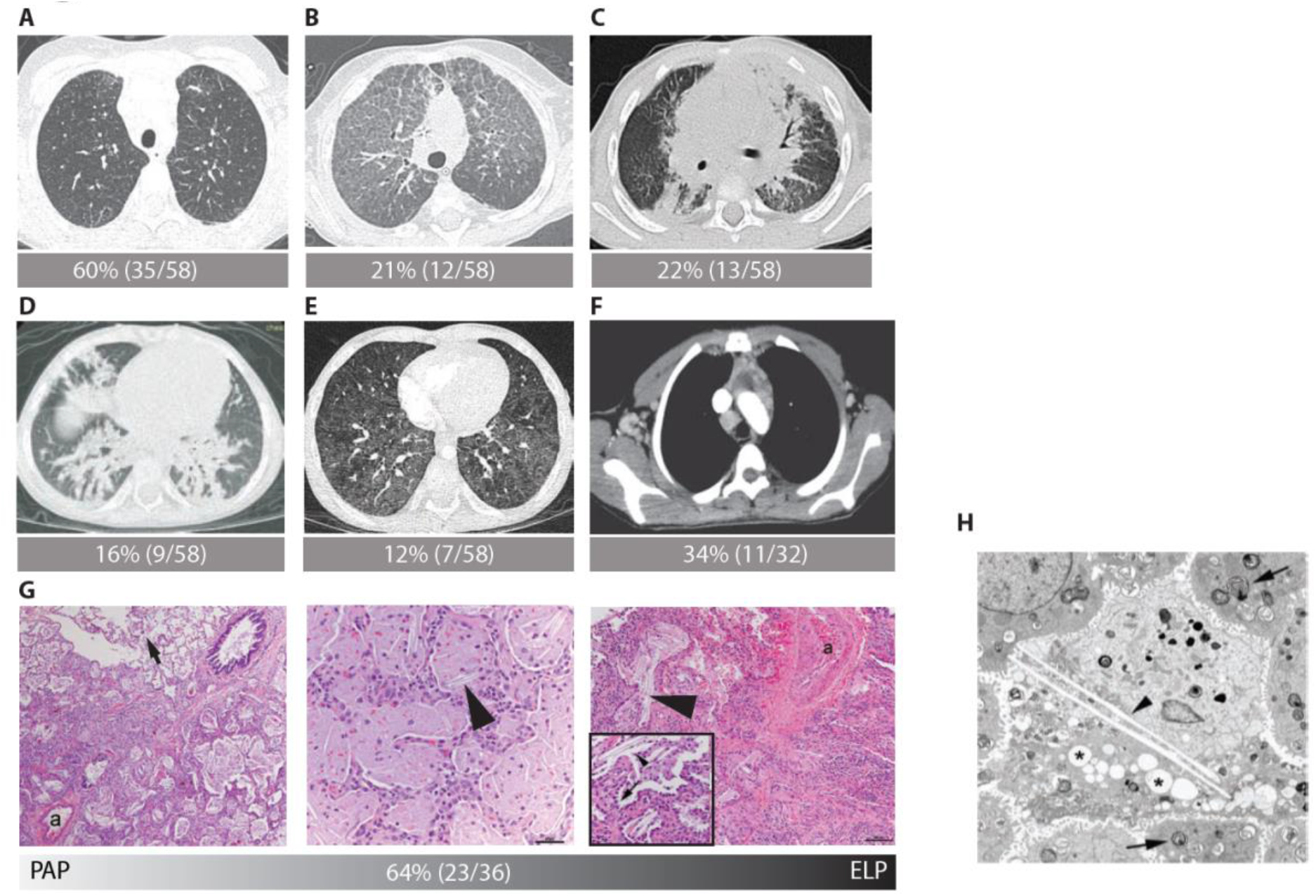
Distinctive radiologic and pathologic features. Panels A-E: Representative axial chest CT images: A. Multilobar, predominantly peripheral septal thickening, most marked in the lower lung, parahilar and/or anterior upper lobes with or without adjacent ground glass opacities. B. Crazy-paving C. Peripheral consolidations. D. Peribronchovascular consolidations. E. Predominantly ground-glass opacities. F. Hyper-enhancing lymph nodes on contrast-enhanced CT. Panels G-H: Histopathologic findings (hematoxylin & eosin staining) along the pulmonary alveolar proteinosis/endogenous lipoid pneumonia (PAP/ELP) spectrum. Alveolar filling with eosinophilic proteinaceous material (**figure 2G left**), admixed with a variable degree of ELP, indicated by cholesterol clefts (arrowheads) and foamy (lipid-containing) macrophages (**figure 2G middle and right**), as described.[76,77] Regions of PAP/ELP accompanied by type II alveolar epithelial cell hyperplasia (**figure 2G, right insert, arrow**), mild to moderate interstitial infiltration by inflammatory cells and lobular remodeling (airspace widening with increased interstitial smooth muscle). Typically, PAP/ELP findings were patchy, with involved areas juxtaposed to normal lung (**figure 2G, left, arrow). Figure 2G, right** shows pulmonary arterial wall thickening; a=artery. In A-G, # cases with pattern/# assessable cases are indicated. H. Electron micrograph showing normal lamellar bodies within type II cells (arrows) and macrophage (center), containing lamellar debris, lipid (*) and cholesterol clefts (arrowhead). Original magnification x 7000. 4 PAP/ELP cases (1 each: ABCA3, CSF2RB variant), stained for surfactant proteins (SP-B, proSP-C, SP-D, ABCA-3, TTF-1), demonstrated robust immunoreactivity (not shown).

### Histopathology and related genetics

Biopsy or autopsy tissues of 36/61 patients were available for centralized analysis (GD). Multi-compartment disease (some combination of alveolar, airway, pleural, vascular alterations) was observed in all cases. Using their primary pattern of injury, 3 subgroups were defined: spectrum of pulmonary alveolar proteinosis/endogenous lipoid pneumonia (PAP/ELP), vasculopathy and other (**table S4**). The pathology typically associated with Pattern A on CT is nonspecific interstitial pneumonia (NSIP).[22,23] Surprisingly, in 21 patients with CT Pattern A who had histology, only 1 had NSIP. Instead, the predominant pathology (64% of cases reviewed) was PAP/ELP (**figure 2G**), which was patchy, and often accompanied by associated vascular changes (**figure 2G, right**). PAP/ELP is very rare in rheumatic disease,[24] and the CT pattern typically associated with PAP is crazy paving (Pattern B).[25]

To identify other histologic features associated with PAP/ELP-like pathology in our cases, we generated a heat map (**figure S5A**). Not surprisingly, type II alveolar cell hyperplasia was highly associated with PAP/ELP.[26,27] The next associated finding was lymphoplasmacytic inflammation (71% of PAP/ELP cases). Third, (55%) was mild to moderate pulmonary arterial wall thickening. Hypertensive vascular changes are not typically associated with inherited PAP/ELP, suggesting a secondary disease process.[26-28] Electron microscopy (available in 9 PAP/ELP cases) demonstrated well-formed lamellar bodies within type II alveolar epithelial cells. Variable accumulation of macrophages containing lamellar debris, lipid and cholesterol clefts was observed (**figure 2H**). The EM findings are more characteristic of macrophage overloading or dysfunction than of genetic disorders in surfactant metabolism.[29,30] When we examined 13 PAP/ELP cases (6 with EM analysis) for genes causing hereditary PAP [*SFTPA1, SFTPB, SFTPC, ABCA3, NKX2-1, CSF2RA, CSFR2B, MARS*],[26,28,31-35] 6 were heterozygous for protein-changing mutations, but none was *de novo* in trio analyses (**table S5A**). One (*SFPTC* p.R167Q) causes PAP with low penetrance;[31] the others are not known to cause PAP in heterozygotes. While these rare variants (maximum allele frequency <5%) might contribute to LD in these children, they are not likely the full explanation.[32,36, 37]

Vascular abnormalities were the predominant finding in 4/36 biopsies (**figure S5B-D, table S4**). Consistent with chest CT findings, interstitial fibrosis was generally mild, with advanced fibrosis/remodeling in only 4/36 samples, including 2 autopsies (**figure S5D)**.

### Candidate risk factors for parenchymal lung disease

#### Other genetic factors

No evidence for a shared monogenic explanation for LD was found in WES of 20 cases analyzed (not shown). We also assessed the frequency of HLH/MAS-related gene variants (*PRF1, LYST, STX11, STXBP2, UNC13D, NRLC4*). Rare protein-altering variants, all heterozygous and none *de novo*, were found (**table S5B**). Concordance between these variants and MAS (at sJIA onset or ever during disease course to data close) was not observed. The frequency of such variants (55%) is higher than reported for sJIA with MAS (36%)[38] and could contribute to propensity for inflammation.

#### Early onset sJIA

Compared to control sJIA subjects in the CR, the LD cohort’s median age at sJIA onset was substantially younger (**table 1:** 2.3y [1.1-5.0] vs. 5.2y [2.8-9.8] p=1⨯10^−7^; **figure S6A**). The CR cohort showed the full age of onset range of published sJIA cohorts, whereas the LD cohort was similar to a subgroup with younger onset age in sJIA cohorts[39,40] (**figure S6B**). Within LD cases, early onset of sJIA/sJIA-like disease was tightly correlated with PAP/ELP-like pathology (Wilcoxson test, p= 2.3e-4; if cut-off for age at sJIA onset <5y, OR = 15, Fisher p=0.001, compared to older children in the cohort). 91% of cases (21/23) with PAP/ELP had sJIA onset at <5y (**figure S6C**).

#### Trisomy 21

In the LD cohort, T21 prevalence (**table 1**) was strikingly higher (10%) than in sJIA registry cohorts [0.2%: 1/471 in the CR and 2/914 in PharmaChild, these being similar to the frequency (0.14%) in live births.[41] There were suggestions of more aggressive LD in these children, all of whom developed LD on anti-IL-1/IL-6. 4/6 children with T21 were hypoxic (OR=7.8, Fisher p=0.08, compared to the proportion of non-T21 with hypoxia). 2/5 with T21 showed advanced interstitial fibrosis/remodeling (OR=8.4, Fisher p=0.09), and 2/4 children with advanced fibrosis had T21. 5/6 (83%) had viral or fungal lung infection at LD diagnosis, compared to 16/55 (29%) of non-T21 (OR=12, Fisher p=0.02)

#### Pre-exposure to cytokine inhibitors

Compared to previously described sJIA patients,[18] the LD cohort demonstrated rare clinical, radiologic and pathologic findings. During the time period of the series, the annual number of cases in the LD cohort increased dramatically, although some bias of ascertainment is possible due to increased awareness of this disease (**figure 3A**). PAP/ELP pathology increased among biopsied cases (**figure 3B**). The proportion of cases exposed to IL-1/IL-6 inhibitors increased in the cohort **(figure 3C)** and in all reported lung disease cases in sJIA (**figure S7**). These 3 trends coincided with increased use of IL-1/IL-6 inhibitors for sJIA.[42] Thus, we asked whether exposure to these inhibitors was related to the prevalence of unusual features. The frequencies of acute clubbing, digital erythema, unexplained abdominal pain, peripheral eosinophilia, CT Pattern A or D, hyper-enhancing lymph nodes and PAP/ELP pathology, but not PH, were substantially higher (p<0.1; FDR<20%) in pre-exposed (exposed before LD diagnosis) versus non-pre-exposed subjects (**figure 4A-B)**. Median time from inhibitor start to LD diagnosis was 1.2 years (IQR 0.7-2.0y, n=46).

**Figure 3:**
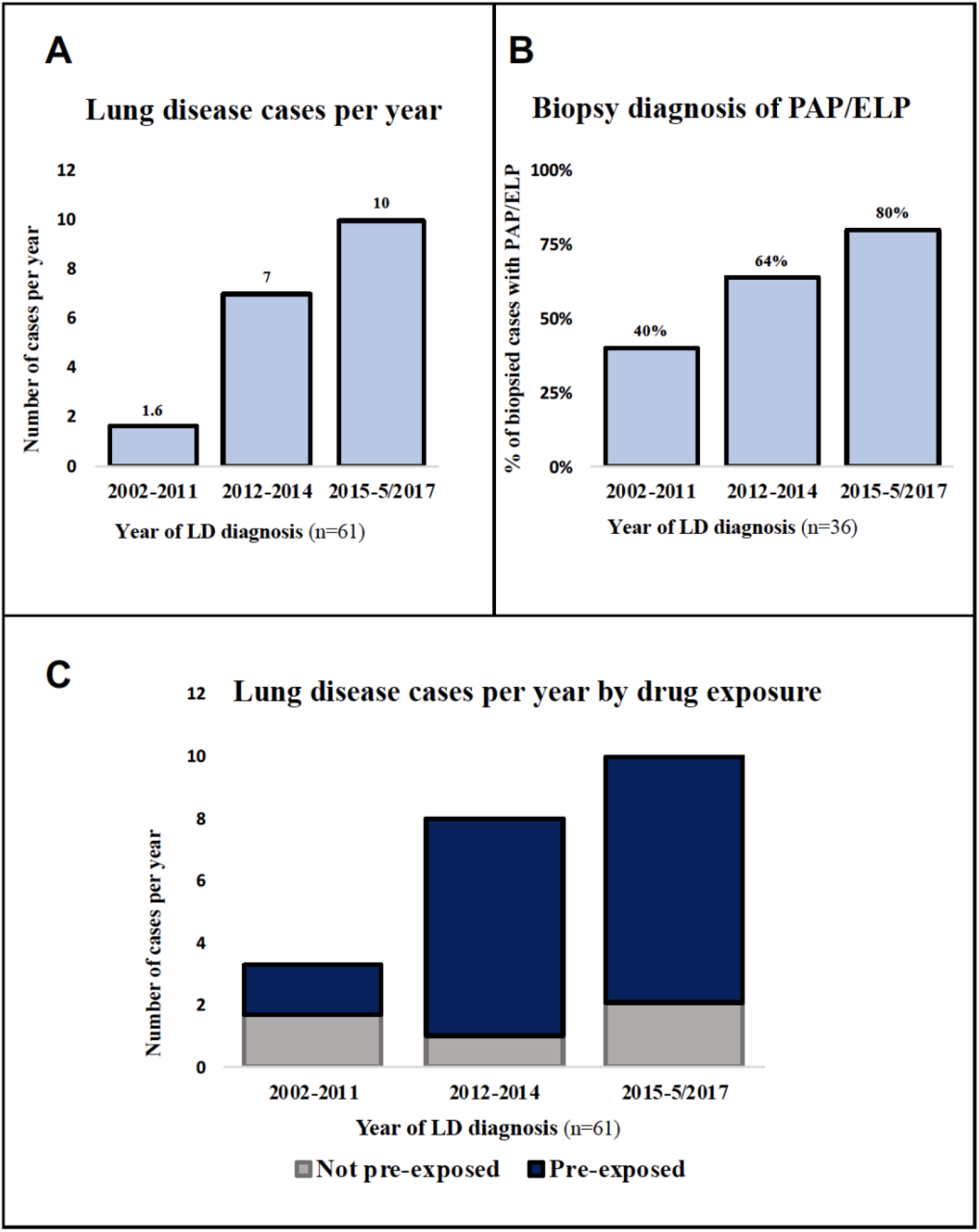
Annual number of reported cases of lung disease (LD) and PAP/ELP pathology. A. Annual number of LD cases in this series (total n=61) B. Percentage of biopsied LD cases (n=36) with PAP/ELP pathology, grouped by year of LD diagnosis C. Annual incidence of LD in our cohort indicating proportions exposed (blue) or not (gray) to anti-IL-1/IL-6 inhibitors are indicated.

**Figure 4:**
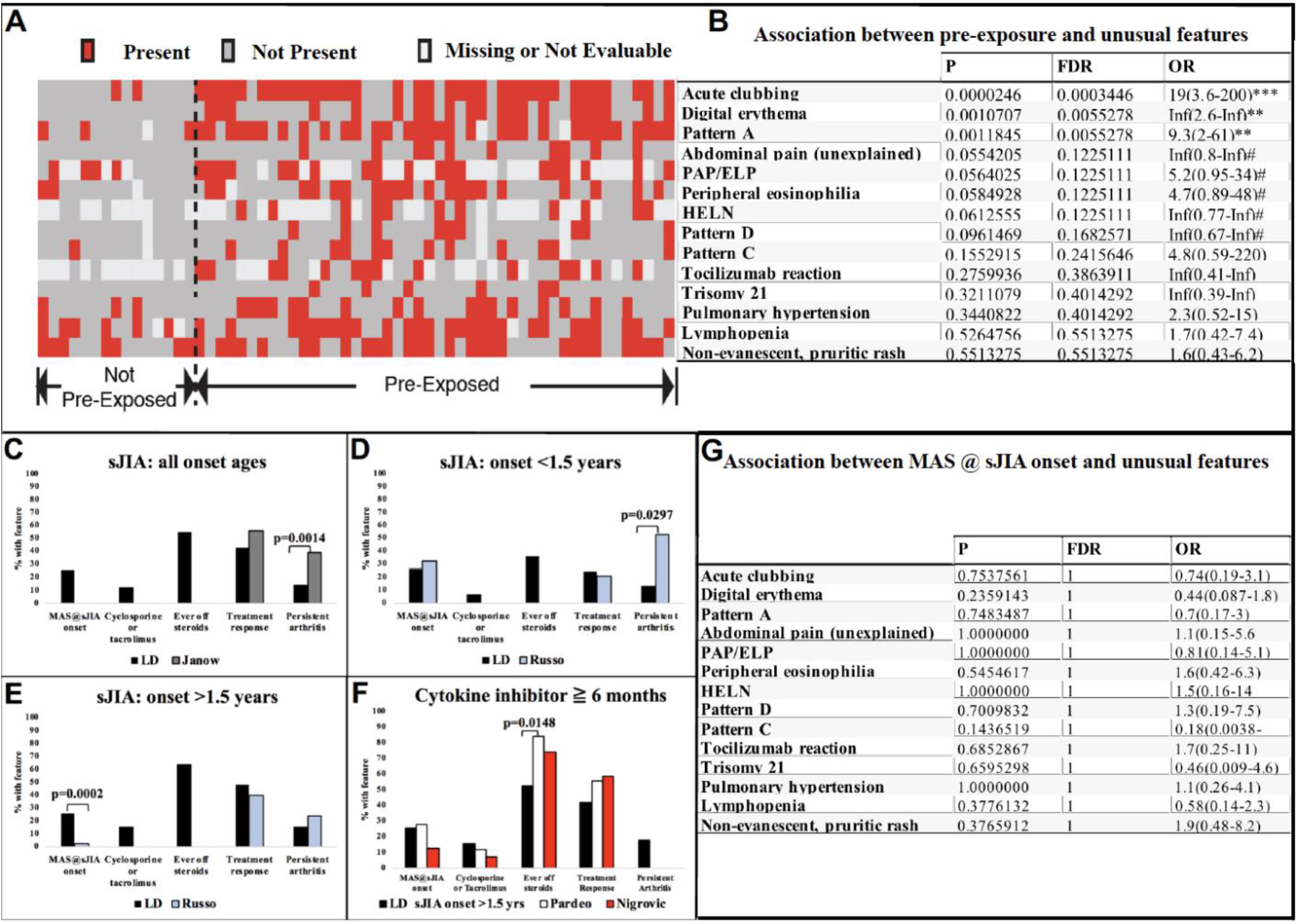
Association between unusual features and pre-exposure to anti-IL-1/IL-6 or MAS at sJIA onset. A. Heat map indicating occurrence of unusual clinical and radiologic features (rows) by subjects (columns), grouped by pre-exposure status. B. Statistical analysis for panel A, indicating P values, false discovery rate (FDR) and odds ratio (OR) with 95% confidence interval. Inf, infinite; #, P < 0.1; *, P < 0.05; **, P < 0.01; *** P < 0.001. C-F. Comparison of severity-related features in pre-exposed LD cases versus published sJIA cohorts. C. Pre-exposed LD cases compared to Janow *et al*.[42] D. Pre-exposed LD cases with sJIA onset <1.5y, compared to comparable age group in Russo *et al*;[43] cut-off at <1.5y was chosen by Russo *et al*, based on developmental difference before versus after 18 months. E. Pre-exposed LD cases with sJIA onset >1.5y, compared to comparable age group in Russo *et al*.[43] F. Pre-exposed LD cases treated with IL-1/IL-6 inhibitors for ≥ 6 mos compared to comparable groups in Pardeo *et al*[78] and Nigrovic *et al*.[79] No bar indicates unavailable data. For details on definitions and published cohorts, see **table S6**. G. Statistical analysis of associations between macrophage activation syndrome (MAS) at sJIA onset and unusual clinical features of LD in sJIA, indicating P values, false discovery rate (FDR) and odds ratio (OR) with 95% confidence interval.

#### Severe or refractory sJIA

It was possible that the observed association between drug exposure and unusual features was attributable to a more inflamed sJIA course in those who develop LD (termed “channel bias”). To investigate this possibility, we assessed 5 clinical features associated with sJIA severity, as there is no validated sJIA severity index. For increased severity, we assessed MAS at sJIA onset, need for calcineurin inhibitors, and persistent arthritis; for reduced severity, we assessed “ever off” steroids and positive treatment response. The small sample of non-exposed subjects prevented us from performing a full analysis within our cohort. We compared the pre-exposed LD subgroup to published cohorts (**figure 4C-F; table S6**). In Russo *et al*.[43], the proportion of early sJIA onset children (<1.5 years) with MAS at sJIA onset was 10x higher than the proportion of later sJIA onset (>1.5 years) children (32% vs. 3%). This difference was interpreted to indicate more severe inflammation in early onset sJIA. Among pre-exposed, early-onset sJIA, LD subjects, the proportion with MAS at onset (27%) was comparable to early-onset subjects in Russo *et al*.[43]. For pre-exposed, later-onset sJIA LD children, a significantly higher proportion had MAS at onset versus the comparable group in Russo *et al*.[43] (26% vs 3%; OR=12.75 p=0.0004) and versus another sJIA cohort, Behrens *et al*.[44] (OR=4.83, p=0.003). These observations raised the possibility that LD was associated with MAS at onset, rather than exposure to the inhibitors. Arguing against this possibility, there was no association in the LD cohort between MAS at onset and any of the unusual features (**figure 4G**). Thus, the associations between exposure to anti-IL-1/IL-6 and the unusual features are unlikely to be confounded by higher frequency of MAS at sJIA onset.

The pre-exposed LD cohort was generally similar to published cohorts for frequency of treatment-responsive disease [lack of calcineurin inhibitor, a period of systemic quiescence or substantially reduced steroid treatment; **figure 4C-F**]. One exception was that a lower proportion of the LD subgroup treated with inhibitors for *≥*6 months was ever off steroids; nonetheless, this proportion was >50% (**figure 4F**). In addition, at data close, 58% of the pre-exposed (18/31) reported inactive sJIA (on medication) despite continuing LD in 94% (17/18). These observations argue that refractory sJIA is not required for LD development or persistence.

### Survival

The period of follow-up after LD diagnosis was variable (median 1.7 yrs; IQR 0.75-3 yrs). Survival was drastically lower in the LD cohort (mortality: 159/1000 person-years; **figure 5**) than in a UK cohort of sJIA patients who required biologic agents (mortality: 3.9/1000 person-years)[45]. The predominant cause of death in our cohort was reported as diffuse lung disease (12/22 deaths), with MAS in 5/12 (**table S7**). Among 75 categorical variables (**table S2-S5**), male sex, hypoxia at initial LD evaluation, predominantly neutrophilic BAL (*≥*40% neutrophils, over 10x>normal)[46,47], but not PH, appeared to associate with shortened survival (**figure S8A-D**). These associations were not significant after adjusting for multiple tests. However, BAL neutrophilia (*≥*50%) has been linked to fatality,[48,49] and 100% in our series (12/12) with this feature were deceased by data close.

**Figure 5:**
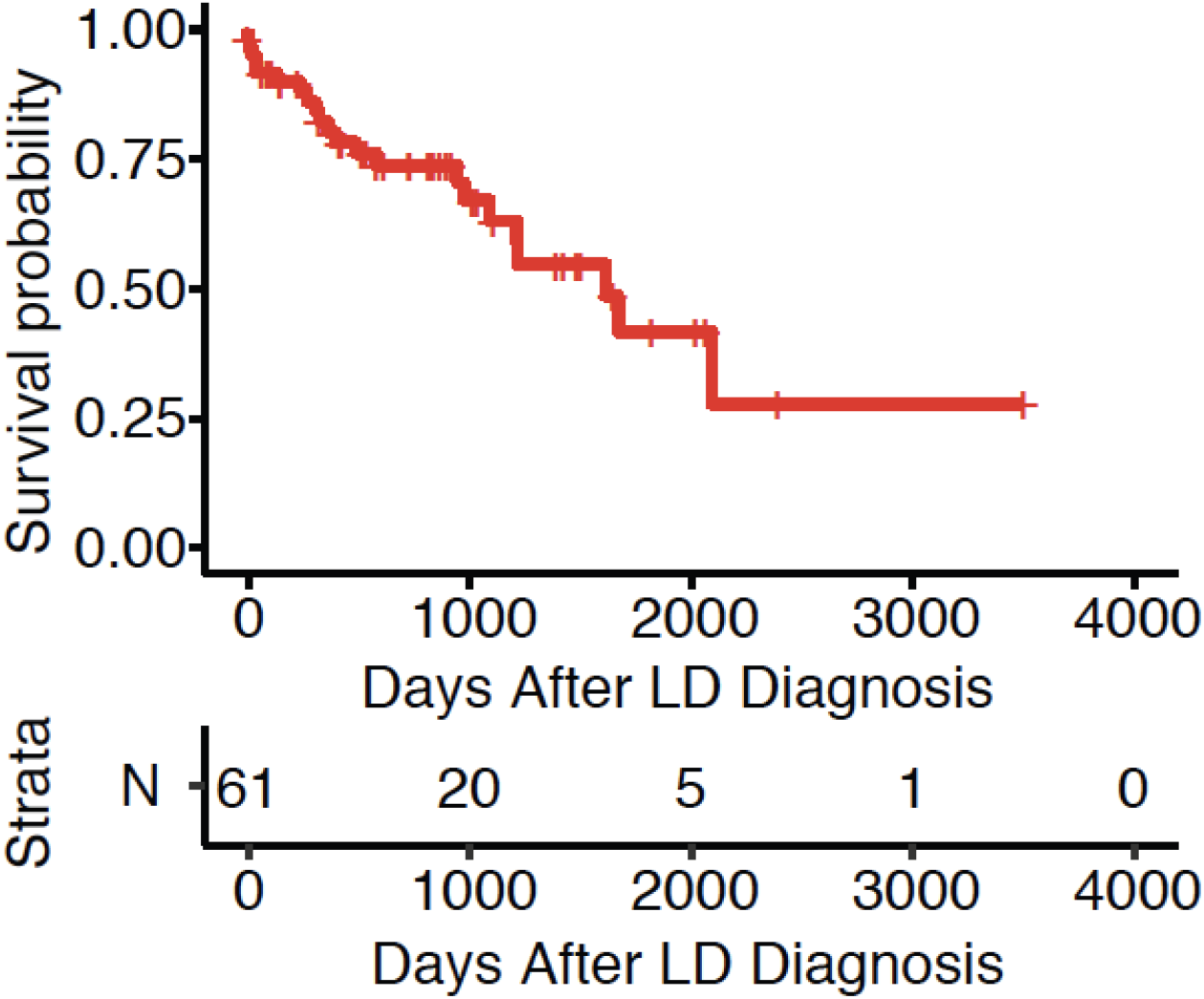
Survival outcome in sJIA cohort with lung disease (LD) The number of survivors at a given time point after lung disease (LD) diagnosis is shown (strata).

## DISCUSSION

LD in sJIA was characterized by young age at sJIA onset and unusual clinical features, including acute erythematous clubbing, atypical rash, and anaphylaxis to tocilizumab; severe tocilizumab reaction in sJIA with pulmonary disease was also noted in data from the PharmaChild registry[50]. The most prevalent finding on chest CT was peripheral septal thickening +/- ground glass opacities. Consolidation, and hyper-enhancing lymph nodes were also observed. On tissue diagnosis, this group showed primarily PAP/ELP-like pathology. Compared to PAP/ELP in other settings, the LD pathology was distinctive for its patchiness and associated vascular changes.

The proportion of LD cases with PAP/ELP-like tissue diagnosis has increased since 2010, coinciding with increasing use of IL-1/IL-6 inhibitors.[42] Pre-exposure to these inhibitors was characteristic of the predominant phenotypic subtype in our series. It is possible that this association is confounded by concomitant reduction in steroids with inhibitor use or by treatment of severe inflammation; our data do not conclusively rule out these possibilities. However, severe disease has been observed since the initial description of sJIA in 1897,[51] whereas the LD with associated features described here appears to be new and increasing in frequency (see below). Among biopsied cases, PAP/ELP was found in 80% with pre-exposure (includes one with mostly pleural sample and limited PAP foci) vs. 36% not pre-exposed (OR=7, (95% CI:1.45-33,7), Fisher p=0.015; **figure S5E**) and was independent of PAP/ELP association with young age (**figure S9**). Thus, IL-1/IL-6 inhibitor exposure may be associated with development of PAP/ELP-like disease in a subset (apparently small) of sJIA patients, among the substantially larger group of patients who derive striking benefit from these inhibitors.

Autopsy RNA sequencing data from the Genotype-Tissue Expression (GTEx) Project show the lung is a major physiologic producer of IL-1 and IL-6 in adults (**figure S10**),[52] and cytokine profiling suggests that circulating IL-1RA levels (reflecting the IL-1 activity) in young (<4y) healthy children are 2x higher than in older healthy children.[53] In addition, NF*κ*B, a key transcription factor downstream of IL-1, stimulates angiogenesis and alveolarization in the post-natal, developing lung.[54] These observations raise the possibility of a physiologic role of IL-1/IL-6 in the lung, particularly in early childhood. The striking enhancement of LD risk by early age of sJIA onset suggests developmental vulnerabilities that may interact with the inhibitors.

A specific relationship between reduced IL-1 and PAP development is described in mice. IL-1*α*^-/-^ mice (but not IL-1*β*^-/-^ mice) challenged with inhaled silicone, an inflammasome (NLRP3) activator,[55] develop PAP-like lung disease.[56] In the lung, IL-1 regulates GM-CSF levels[57,58] and macrophage function;[56,57] disruption of either can lead to surfactant accumulation and PAP.[59,60] These findings imply a link between reduced IL-1 and PAP, but also suggest that additional triggers may be required for disease development, in line with the rarity of severe parenchymal LD among the overall population of sJIA patients treated with inhibitors.

We found an outsized risk of LD in children with T21 and sJIA. T21 carries increased susceptibility to adverse drug reactions[61] and to viral pneumonia.[62] Another contributing factor may be underlying type 1 interferonopathy, recently described in T21.[63]

A subset of LD cases met criteria for DReSS, a delayed form of severe drug-related hypersensitivity with organ involvement that can include lung [17,64,65](**table S8**). DReSS findings included dramatic eosinophilia, often despite concurrent steroids, together with extensive, persistent rash, frequently involving the face, which is uncommon in sJIA.[18] Altered drug metabolism in childhood may increase risk of hypersensitivity reactions.[66]. Another consideration is drug-induced interstitial lung disease (DiILD), previously reported in children with rheumatic disease on biologics.[67] DiILD has overlapping chest CT findings with LD in sJIA.[68] No pathologic findings are pathognomonic of DiILD, [68,69] but PAP has been described.[24]

Infection may exacerbate LD and may trigger its detection. Pathogens identified at initial lung evaluation (**table S3**) included rhinovirus (a cause of severe lower respiratory infection in young children[70]), herpes viruses and pneumocystis, all of which require IL-1 for optimal host defense.[55,71,72] Pneumocystis pneumonia (PCP) is a recognized cause of PAP[25] and is associated with high mortality in immunocompromised individuals.[73] PCP risk is also elevated in DReSS.[74] At least 4 of our cases had PCP; these were diagnosed by PCR of BAL, the preferred test in non-HIV immunosuppressed patients.[73] It seems prudent to consider prophylaxis for sJIA patients with lymphopenia or steroid use (consistent with recommendations[75]) or T21.

In 23/61 cases, LD detection occurred with concurrent MAS. 19/61 had their first MAS episode at or after LD diagnosis, suggesting that LD may trigger systemic inflammation. Pulmonary hypertension (PH) with a range of severity was observed, with or without pre-exposure to inhibitors and with or without PAP/ELP pathology. 2/20 subjects lacked substantial parenchymal lung disease at PH detection. Together, these observations argue that PH in this cohort reflects heterogenous biology.

Over 50 additional LD cases in sJIA have been reported to us since closing this series. The FDA adverse event website (FAERS) shows 39 adults [rheumatoid arthritis (23); AOSD (11); other (5)] developing alveolar disease or PH on IL-1/IL-6 inhibitors and 4 DRESS cases (second quarter, 2019). The associations between cytokine inhibition and LD in sJIA and the mechanistic hypotheses regarding these observations demand further investigation. Importantly, we acknowledge the limitations of retrospective data, use of historical/published data for controls, possible biases as mentioned, and false discovery issues associated with multiple hypothesis testing and exploration of data in a large data set. With these limitations, one cannot assign causality to cytokine inhibition in LD in sJIA.

Likewise, it is premature to make treatment recommendations solely on the basis of our findings. Therapeutic decision making is challenging in the setting of sJIA-associated LD, particularly given the efficacy of the inhibitors in sJIA/sJIA-like disease [4-6]. Management strategies should continue to be devised on an individual basis for these patients. However, in light of the high fatality with current treatment approaches, efforts to determine lung disease prevalence, uncover molecular mechanism(s) and devise treatment and prevention approaches are urgently needed.

## Data Availability

Data are available upon reasonable request.

## ACKNOWLEDGMENTS

The CARRA registry has been supported by The NIH’s National Institute of Arthritis and Musculoskeletal and Skin Diseases (NIAMS) & the Arthritis Foundation. We thank all participants and hospital sites that recruited patients for the CARRA Registry.

## CARRA Registry site principal investigators and research coordinators

K. Abulaban, R. Agbayani, S. Akoghlanian, E. Anderson, M. Andrew, B. Badwal, L. Barillas-Arias, K. Baszis, M. Becker, H. Bell-Brunson, H. Benham, S. Benseler, T. Beukelman, J. Birmingham, M. Boncek, H. Brunner, A. Bryson, H. Bukulmez, L. Cerracchio, E. Chalom, J. Chang, N. Chowdhury, K. Chundru, T. Davis, J. Dean, F. Dedeoglu, V. Dempsey, M. Dionizovik-Dimanovski, L. Dolinsky, J. Drew, B. Feldman, P. Ferguson, B. Ferreira, C. Fleming, L. Franco, I. Goh, D. Goldsmith, B. Gottlieb, T. Graham, T. Griffin, M. Hance, D. Helfrich, K. Hickey, M. Hollander, J. Hsu, A. Huber, A. Hudson, C. Hung, A. Huttenlocher, L. Imundo, C. Inman, J. Jaquith, R. Jerath, J. Jones, S. Jones, L. Jung, P. Kahn, D. Kingsbury, K. Klein, M. Klein-Gitelman, S. Kramer, A. Kufen, S. Lapidus, D. Latham, S. Linehan, B. Malla, M. Malloy, A. Martyniuk, T. Mason, K. McConnell, D. McCurdy, K. McKibben, C. McMullen-Jackson, K. Moore, L. Moorthy, E. Muscal, W. Norris, J. Olson, K. O’Neil, K. Onel, K. Phillips, L. Ponder, S. Prahalad, C. Rabinovich, S. Rauch, S. Ringold, M. Riordan, S. Roberson, A. Robinson, E. Rojas, M. Rosenkranz, B. Rosolowski, N. Ruth, K. Schikler, A. Sepulveda, C. Smith, H. Stapp, K. Stewart, R. Syed, A. Tangarone, M. Tesher, A. Thatayatikom, R. Vehe, E. von Scheven, D. Wahezi, C. Wang, C. Wassink, M. Watson, A. Watts, J. Weiss, P. Weiss, A. Wolverton, J. Woo, A. Yalcindag, Q. Yu, A. Zeft, L. Zemel, A. Zhu, J. Zwerling.

The authors thank Drs. Joseph A. Kovacs, Jay K. Kolls and Sergio Vargas for their expert input on pneumocystis pneumonia, Dr. James Verbsky for contributing unidentified genetic data, Dr. Yuki Kimura for providing de-identified diagnosis dates from her study and Dr. Xuan Qin for staining available tissue samples for PCP. The authors also thank Dr. Claudia Macaubas for statistical assistance and Ms. Melinda Hing for assistance with the manuscript.

## Contributors

VS, GC, GD, RPG, AL, JB, KJ, JX, RB, LB, YL, LT, TD, PK, EM: collection, analysis, discussion and interpretation of data. VS, GC wrote the manuscript. EM, PK: checked and revised the manuscript. SC, GS, RD, XQ, KA, KB, EB, JB, AC, MC, RC, AD, FD, BG, AG, IF, MF, SG, LY, MS, AH, KH, MH, LH, MI, CI, RJ, KK, DK, MK-G, KL, SL, CL, JL, DL, DM, JM, KO, SO, MP, KP, SP, SR, AR, MR, NR, JR, RS, DS, SS, JS, HS, CT, SV, RV, JY, GB, MD: provided data, checked and approved the manuscript.

## Patient and public involvement statement

Patients were not involved in the research process of this study. Results will be shared via CARRA patient communication mechanisms.

## Patient consent for publication

Not applicable

## Ethics approval

Ethics approval was obtained through the Stanford University School of Medicine institutional review board. Contributing case reporters obtained approval per local institutional review board requirements.

## Public/Private

Not applicable

## Data availability statement

Data are available upon reasonable request.

